# Prognostic Value of Long Noncoding RNA NORAD in Various cancers: a meta-analysis

**DOI:** 10.1101/2020.07.09.20150185

**Authors:** Qin Yang, Zheng Zhang, Yuan-Yuan Gong, Zhi-Ran Li, Hua-Zhu Zhang, Gong-Hao He

## Abstract

**Objective:** Accumulating studies reported that noncoding RNA activated by DNA damage (NORAD) was correlated with poor survival outcomes for patients in different cancers. However, the effects of NORAD on cancer prognosis were controversial. Therefore, a meta-analysis was carried out to elucidate this issue.

**Methods:** Literature search was performed to collect eligible relevant publications until June 2020. The pooled hazard ratios (HRs) or odds ratios (ORs) with 95% confidence intervals (CIs) were calculated to assess the association of NORAD with prognosis and clinical features in diverse cancers. In addition, bioinformatics analysis was also utilized to validate the results of the meta-analysis.

**Results:** Fourteen relevant articles involving 867 patients were enrolled in the present study. The pooled results showed that elevated expression of NORAD was a risk factor for overall survival (HR = 1.46, 95% CI: 1.06-2.01, P = 0.020), disease-free survival (HR = 1.74, 95% CI: 1.18-2.57, P = 0.005) and recurrence-free survival. Besides, overexpression of NORAD significantly correlated with lymph node metastasis and T stage. Additionally, bioinformatics analysis further strengthened and complemented the results of the present study.

**Conclusion:** Our results showed that NORAD was a risk factor for survival outcomes and clinicopathological parameters in cancer patients. These findings indicated that NORAD may be a promising candidate for prognosis prediction and potential therapeutic target in diverse cancers.

## 1 Introduction

Cancer is a life-threatening disease with high mortality worldwide [1]. It was well acknowledged that early diagnosis and treatments of cancer played important roles in reducing cancer-related mortality rate [2]. However, partly due to the lack of appropriate sensitivity biomarkers for early stage diagnosis and effective treatments, the prognosis and long-term survival rate for various cancers still remained poor and low so far [3,4] and identifying promising sensitive cancer-related biomarkers for early diagnosis, precise treatment and prognosis prediction would be an unmet medical need in current situation.

Long non-coding RNAs (lncRNAs), a class of RNAs without protein-coding ability [5], were actively participated in cell differentiation and proliferation, and thus the aberrant expression of lncRNAs were relevant to diverse diseases, especially cancers [6-8]. Increasing reports revealed that dysregulation of lncRNAs were correlated with the pathogenesis of cancer [9-12]. Furthermore, differentially expressed lncRNAs were also demonstrated to be implicated in the progression of cancers (i.e., invasion, metastases and apoptosis) [5,13-14]. Besides, recent accumulating studies demonstrated that many cancer-related lncRNAs were correlated with the diagnosis and prognosis of cancer patients and might be potential targets for cancer therapy [15-17]. Therefore, lncRNAs were increasingly regarded as promising biomarkers for cancer diagnosis, prognosis and therapies.

Noncoding RNA activated by DNA damage (NORAD, also known as LINC00657) is a kind of lncRNAs acting as vital regulators in the development of cancers [18]. Accumulating previous studies indicated that NORAD expression level was a poor prognostic factor for patients in different cancers (e.g., cervical cancer, bladder cancer, breast cancer) [19-21]. However, individual studies regarding the correlation of NORAD expression with prognosis provided controversial results. For instance, it was reported that high NORAD expression could predict poor prognosis in hepatocellular cancer (HCC) [22], while another study indicated that patients with low expression of NORAD was prone to poor survival [23]. In addition, studies regarding the correlation between NORAD and clinical features, such as tumor size [24,25], lymph node metastasis [26,27] and distant metastasis [25,27], were also inconsistent. Furthermore, owing to relatively small sample size in those previous studies, consensus about the prognostic and clinical role of NORAD in cancers was not reached yet. Therefore, the effect of NORAD expression on prognosis and clinicopathological features of cancer patients is still unclear and a more comprehensive meta-analysis should be carried out to elucidate this issue.

Based on the abovementioned background, a quantitative meta-analysis was herein carried out to explore the prognostic and clinical significance of NORAD in diverse cancers, hoping to provide a comprehensive insight into the prognostic significance of NORAD in cancer patients.

## 2 Material and Methods

### 2.1 Literature retrieval

A literature search was carried out to obtain eligible publications using PubMed, EMBASE, Cochrane library, China National Knowledge Infrastructure, Wan Fang Database and VIP database until June 2020. We used the following terms for searching: “NORAD long non-coding RNA” or “long non-coding RNA NORAD” or “NORAD lncRNA” or “LINC00657”, “tumor” or “neoplasms” or “cancer” or “malignancy”. Details regarding the search strategy for PubMed were shown in Supplementary Table 1. Moreover, the references of included literatures were also retrieved to identify potentially related articles. In addition, languages were limited to English and Chinese.

**Table 1.** Characteristic of articles included meta-analysis

| Study | Year | Country | Cancer type | Sample | Criterion of high expression | Detection method | Outcome measures | Follow up ( month ) | Date extraction method | NOS |
| --- | --- | --- | --- | --- | --- | --- | --- | --- | --- | --- |
| Hu | 2017 | China | HCC | 49 | Median value | qRT-PCR | OS | 48 | K-M | 7 |
| Li | 2017 | China | PDAC | 33 | NR | qRT-PCR | OS RFS | 45 | K-M | 7 |
| Wu | 2017 | China | ESCC | 106 | Median value | qRT-PCR | OS | 60 | Report | 8 |
| Huo | 2018 | China | CC | 47 | NR | qRT-PCR | OS | 60 | K-M | 7 |
| Zhang | 2018 | China | CRC | 47 | Median value | qRT-PCR | OS | 60 | K-M | 7 |
| Lei | 2018 | China | Colon cancer | 80 | NR | qRT-PCR | OS | 168 | K-M | 8 |
| Li | 2018 | China | BC | 90 | Mean value | RT-qPCR | OS | 60 | Report | 9 |
| Chu | 2019 | China | GBM | 40 | Mean value | RT-qPCR | OS DFS | 24 | K-M | 7 |
| Miao | 2019 | China | GC | 111 | Median value | qRT-PCR | OS | 60 | K-M | 8 |
| Zhou | 2019 | China | Breast cancer | 21 | NR | Q-PCR | OS | 60 | K-M | 8 |
| Yang | 2019 | China | HCC | 95 | NR | qPCR | OS DFS | 100 | Report | 9 |
| Bi | 2020 | China | PDAC | 56 | NR | RT-qPCR | OS | 60 | K-M | 6 |
| Huang | 2020 | China | NSCLC | 60 | NR | RT-qPCR | OS | 60 | K-M | 7 |
| Xu | 2020 | China | OSCC | 32 | NR | qRT-PCR | OS | 60 | K-M | 6 |

GC: gastric cancer; HCC: hepatocellular carcinoma; CRC: colorectal cancer; ESCC: Esophageal squamous cell carcinoma; OSCC: Oral squamous cell carcinoma.
CC: Cervical cancer; BC: Bladder cancer; GBM: Glioblastoma; PDAC: Pancreatic ductal adenocarcinoma; NSCLC: Non-small-cell lung cancer.

### 2.2 Literature selection criteria

The following criteria were applied to include relevant articles: (a) studies focusing on the correlations of NORAD expression with patient survival and clinical parameters in any cancer types; (b) studies dividing subjects into high and low expression groups based on the NORAD expression; (c) studies providing sufficient data to evaluate hazard ratios (HRs) with 95% confidence intervals (CIs) for OS, recurrence-free survival (RFS) and disease-free survival (DFS); (d) full text article was available. Reviews, non-human research, repeated literatures, letters, studies focused on polymorphism and data extracted from TCGA or GEO database were excluded.

### 2.3 Data collection

Two investigators extracted the appropriate data from eligible publications independently. Any disagreements were resolved through discussion or by consulting a third investigator. Information were collected as follows: (a) the basic characteristics of eligible studies, including first author, the date of publication, type of cancer, country, sample size, follow-up data and criteria of high expression, (b) clinicopathological features (i.e., gender, tumor size, tumor number, age, lymph node metastasis, T stage, distant metastasis, liver cirrhosis, differentiation and vascular invasion), and (c) HR and 95% CI for OS, DFS, RFS. If these data were not directly provided in articles, the survival data were extracted from Kaplan–Meier curves using Engauge Digitizer 4.1 and published method [28] was employed to evaluate HR and 95%CI.

### 2.4 Quality assessment

The Newcastle-Ottawa Scale (NOS) was employed to evaluate the quality of included studies and studies with score ≥ 6 were considered as high-quality papers [29].

### 2.5 Bioinformatics analysis

Gene Expression Profiling Interactive Analysis (GEPIA) was utilized to detect the expression of NORAD in different cancer types and to investigate its correlations with OS and DFS based on The Cancer Genome Atlas (TCGA) and the Genotype-Tissue Expression (GTEx) data [30,31]. Besides, correlation analysis was applied between gene expression levels based on GEPIA [30].

### 2.6 Statistical analysis

Pooled HRs and 95% CI were utilized to evaluate the relationship between NORAD expression and clinical outcomes of patients with cancer. ORs and 95% CIs were employed to calculate the association of NORAD expression with clinical characteristics. P < 0.05 was regarded as significant. Heterogeneity among included articles was computed by Q-test and I^2^ index [32]. In case of a significant heterogeneity, the random-effect model was applied (P < 0.05 or I^2^ > 50%). Otherwise, the fixed-effect model was performed. Additionally, sensitivity analysis was carried out to assess the stability of the results by sequentially removing each single study. Furthermore, Begg’s funnel plot and Egger’s test were employed to evaluate the publication bias. STATA 12.0 software was applied to perform all statistical analysis.

## 3 Results

### 3.1 Study characteristics

The literature retrieval identified 171 publications initially, and 29 duplicate literatures and 126 irrelevant articles were excluded after scanning title and abstract. Eventually, 14 literatures were included through detailed assessment (Figure 1).

**Figure 1.**
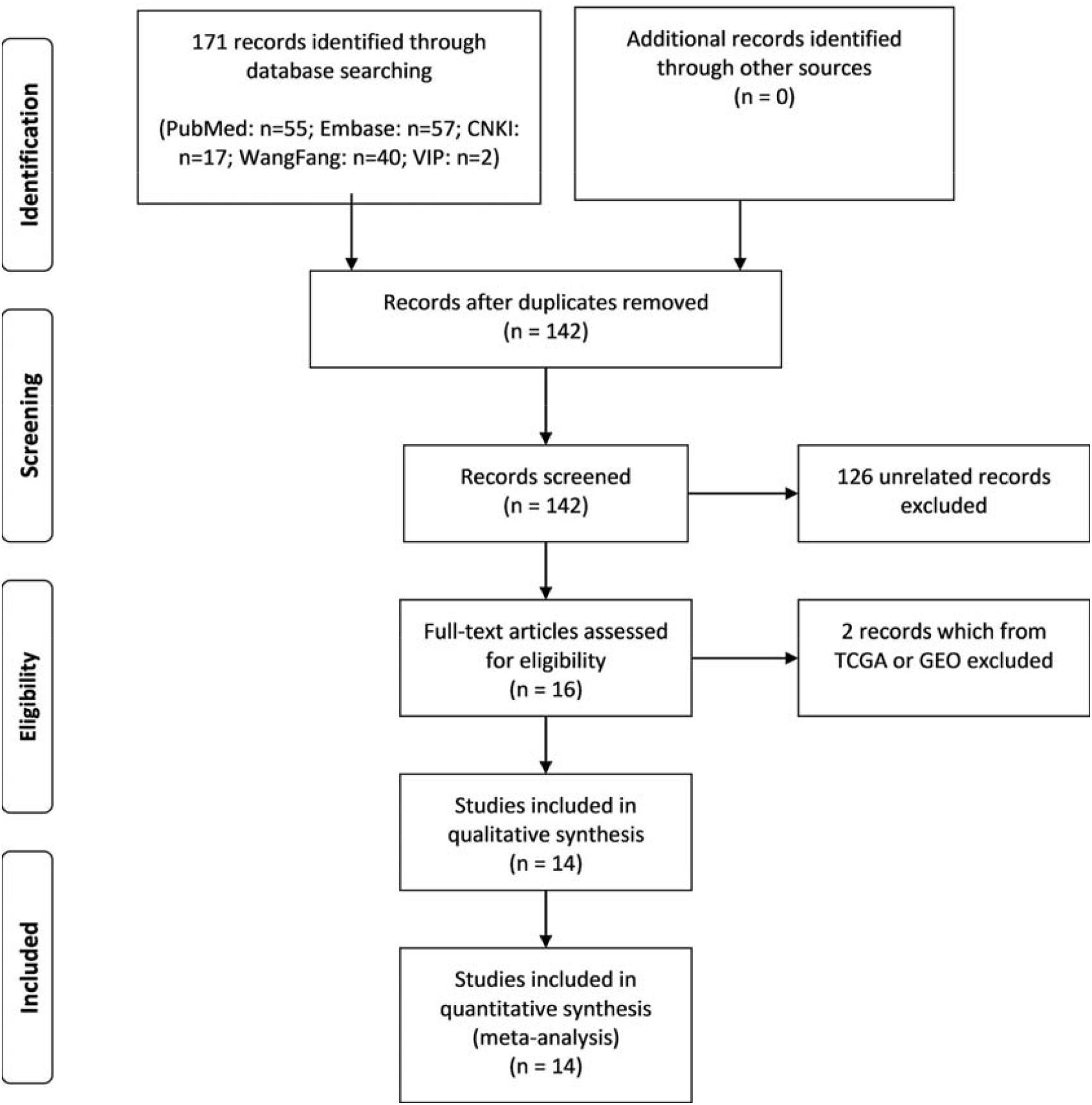
Flow diagram of studies selection process.

Detailed information of included studies was displayed in Table 1. The date of publication ranged from 2017 to 2020. Of these studies, 9 reported clinical features, 14 focused on OS, 2 related to DFS, and only 1 study covered RFS. In addition, the current study contains 11 types of cancers, including gastric cancer (GC), HCC, non-small cell lung cancer (NSCLC), breast cancer, colorectal cancer (CRC), pancreatic ductal adenocarcinoma (PDAC), cervical cancer (CC), bladder cancer (BC), glioblastoma (GBM), oral squamous cell cancer (OSCC) and esophageal squamous cell cancer (ESCC). Additionally, all included articles were considered as high quality with the score ranged from 6 to 9.

### 3.2 Correlation of NORAD expression with clinical outcomes

For the correlation of NORAD expression with OS in cancer patients, a total of 14 articles with 867 subjects were enrolled in the present analysis [19-27,33-37]. The overall results showed that elevated level of NORAD was significantly related to worse prognosis (HR = 1.46, 95% CI: 1.06-2.01, P = 0.020) (Figure 2). Due to the relatively high heterogeneity (I^2^ = 63.1%), sensitivity analysis was conducted. The results showed that the association of NORAD expression with OS was not significantly affected by sequentially removing each single study (Figure 3). Furthermore, subgroup analyses based on follow-up time (more or less than 60 months), sample size (more or less than 60 participants) and cancer type were performed to further explore the correlation of NORAD expression with OS. The pooled HRs demonstrated that elevated expression of NORAD was significantly related with poor OS in studies with follow-up ≥ 60 months (HR = 1.70, 95% CI: 1.26-2.30, P = 0.001), while the correlation was not significant with follow-up < 60 months (Table 2). Besides, after stratification by sample size, a significant relationship was observed in studies with sample size > 60 (HR = 1.67, 95% CI: 1.04-2.68, P = 0.034), but not in studies with sample size ≤ 60 (Table 2). As for the cancer types, the pooled results indicated that elevated level of NORAD could estimate poor outcome in NSCLC (HR = 2.15, 95% CI: 1.06-4.35, P = 0.033), GC (HR = 3.00, 95% CI: 1.53-5.88, P = 0.001), CC (HR = 2.69, 95% CI: 1.13-6.41, P = 0.026), BC (HR = 1.30, 95% CI: 1.05-1.61, P = 0.015) and ESCC (HR = 3.42, 95% CI: 1.75-6.70, P = 0.000), while we observed that low expression of NORAD was significantly associated with poor OS in GBM (HR = 0.23, 95% CI: 0.06-0.93, P = 0.040) (Table 2).

**Table 2.**
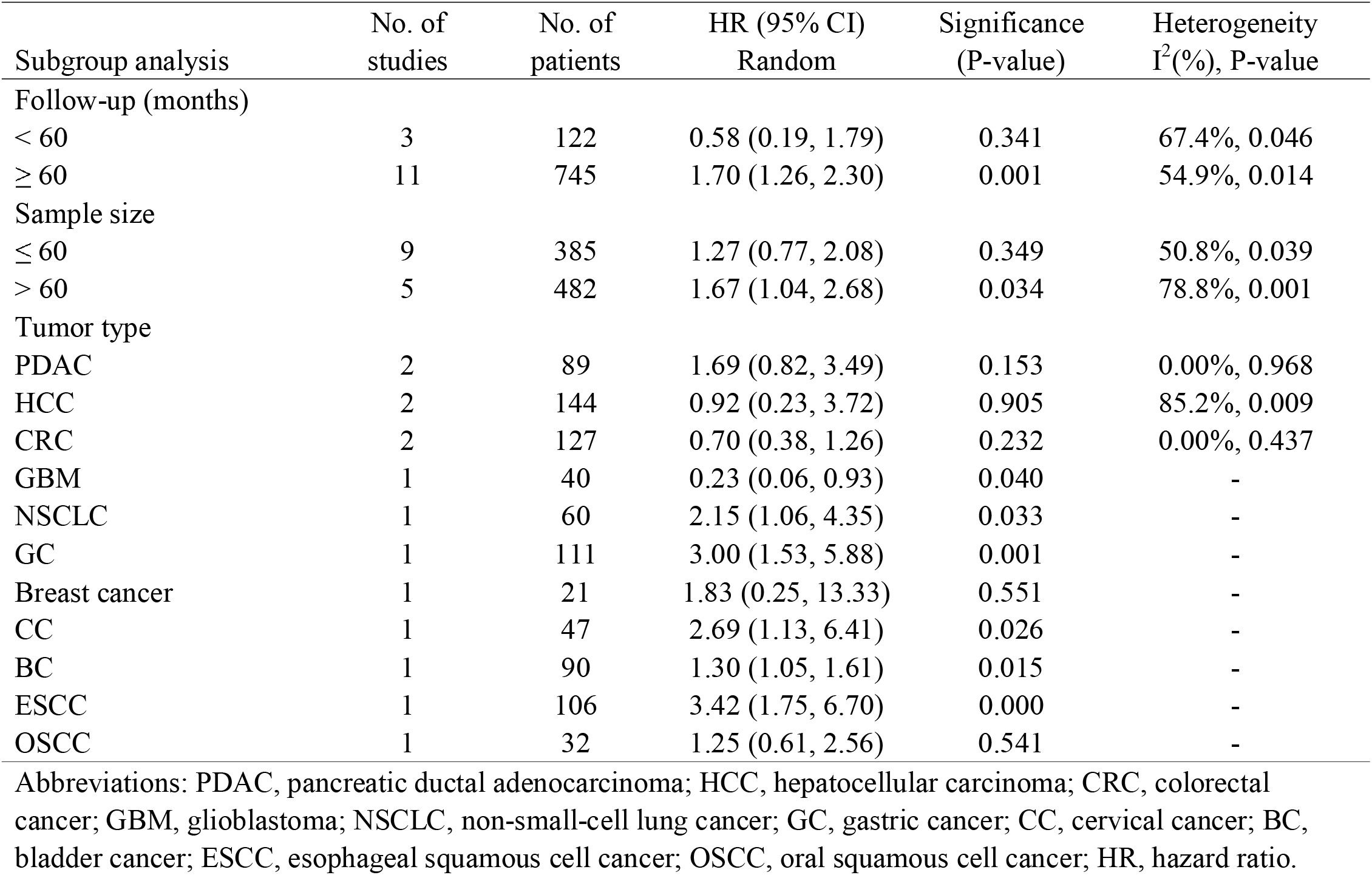
Subgroup meta□analysis of pooled hazard ratios for overall survival

| Subgroup analysis | No. of studies | No. of patients | HR (95% CI)<br>Random | Significance<br>(P-value) | Heterogeneity<br>$I^2$(%), P-value |
| --- | --- | --- | --- | --- | --- |
| Follow-up (months) |  |  |  |  |  |
| < 60 | 3 | 122 | 0.58 (0.19, 1.79) | 0.341 | 67.4%, 0.046 |
| ≥ 60 | 11 | 745 | 1.70 (1.26, 2.30) | 0.001 | 54.9%, 0.014 |
| Sample size |  |  |  |  |  |
| ≤ 60 | 9 | 385 | 1.27 (0.77, 2.08) | 0.349 | 50.8%, 0.039 |
| > 60 | 5 | 482 | 1.67 (1.04, 2.68) | 0.034 | 78.8%, 0.001 |
| Tumor type |  |  |  |  |  |
| PDAC | 2 | 89 | 1.69 (0.82, 3.49) | 0.153 | 0.00%, 0.968 |
| HCC | 2 | 144 | 0.92 (0.23, 3.72) | 0.905 | 85.2%, 0.009 |
| CRC | 2 | 127 | 0.70 (0.38, 1.26) | 0.232 | 0.00%, 0.437 |
| GBM | 1 | 40 | 0.23 (0.06, 0.93) | 0.040 | - |
| NSCLC | 1 | 60 | 2.15 (1.06, 4.35) | 0.033 | - |
| GC | 1 | 111 | 3.00 (1.53, 5.88) | 0.001 | - |
| Breast cancer | 1 | 21 | 1.83 (0.25, 13.33) | 0.551 | - |
| CC | 1 | 47 | 2.69 (1.13, 6.41) | 0.026 | - |
| BC | 1 | 90 | 1.30 (1.05, 1.61) | 0.015 | - |
| ESCC | 1 | 106 | 3.42 (1.75, 6.70) | 0.000 | - |
| OSCC | 1 | 32 | 1.25 (0.61, 2.56) | 0.541 | - |

Abbreviations: PDAC, pancreatic ductal adenocarcinoma; HCC, hepatocellular carcinoma; CRC, colorectal cancer; GBM, glioblastoma; NSCLC, non-small-cell lung cancer; GC, gastric cancer; CC, cervical cancer; BC, bladder cancer; ESCC, esophageal squamous cell cancer; OSCC, oral squamous cell cancer; HR, hazard ratio.

**Figure 2.**
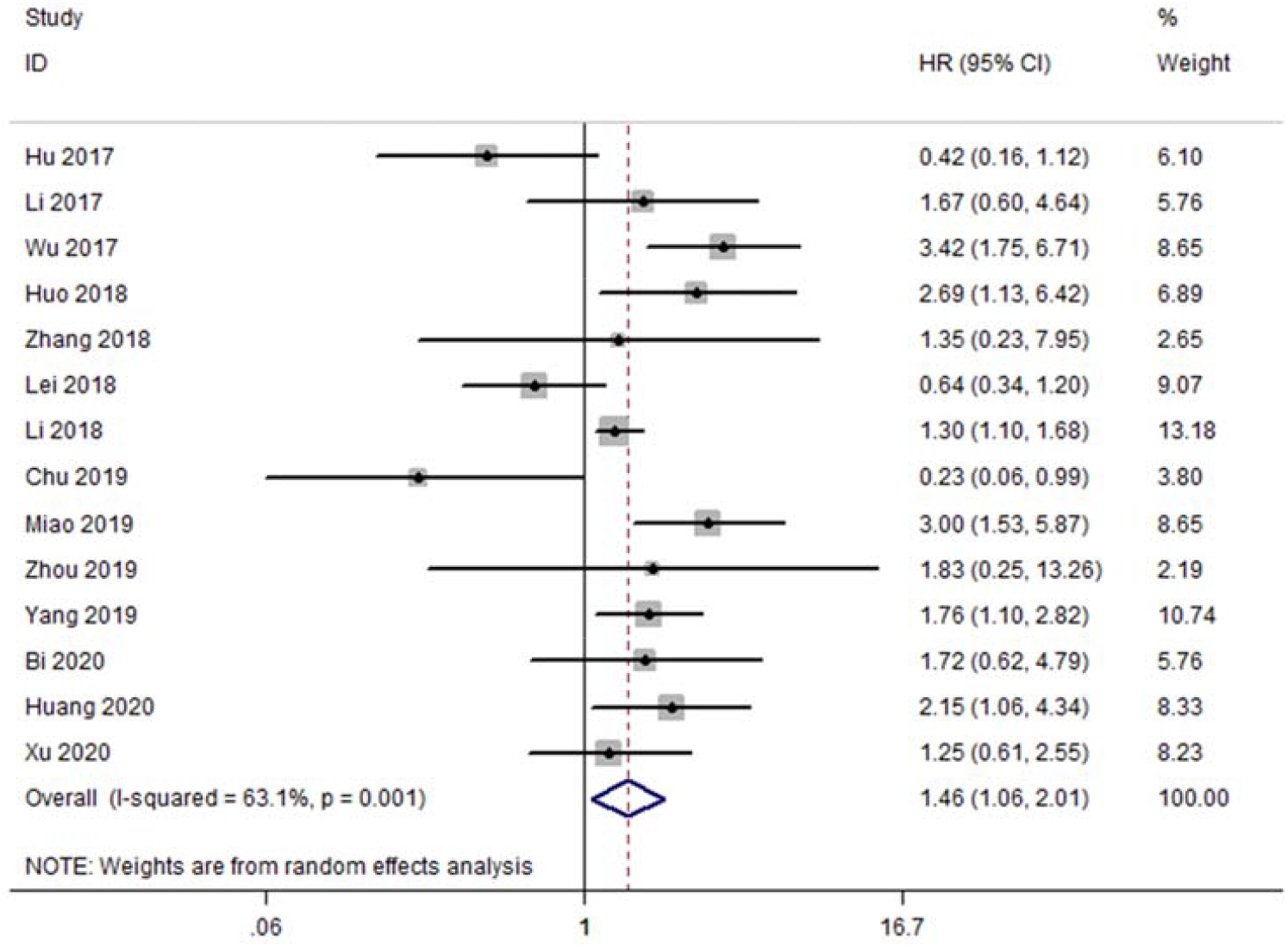
Forest plot for the association between NORAD expression levels and OS.

**Figure 3.**
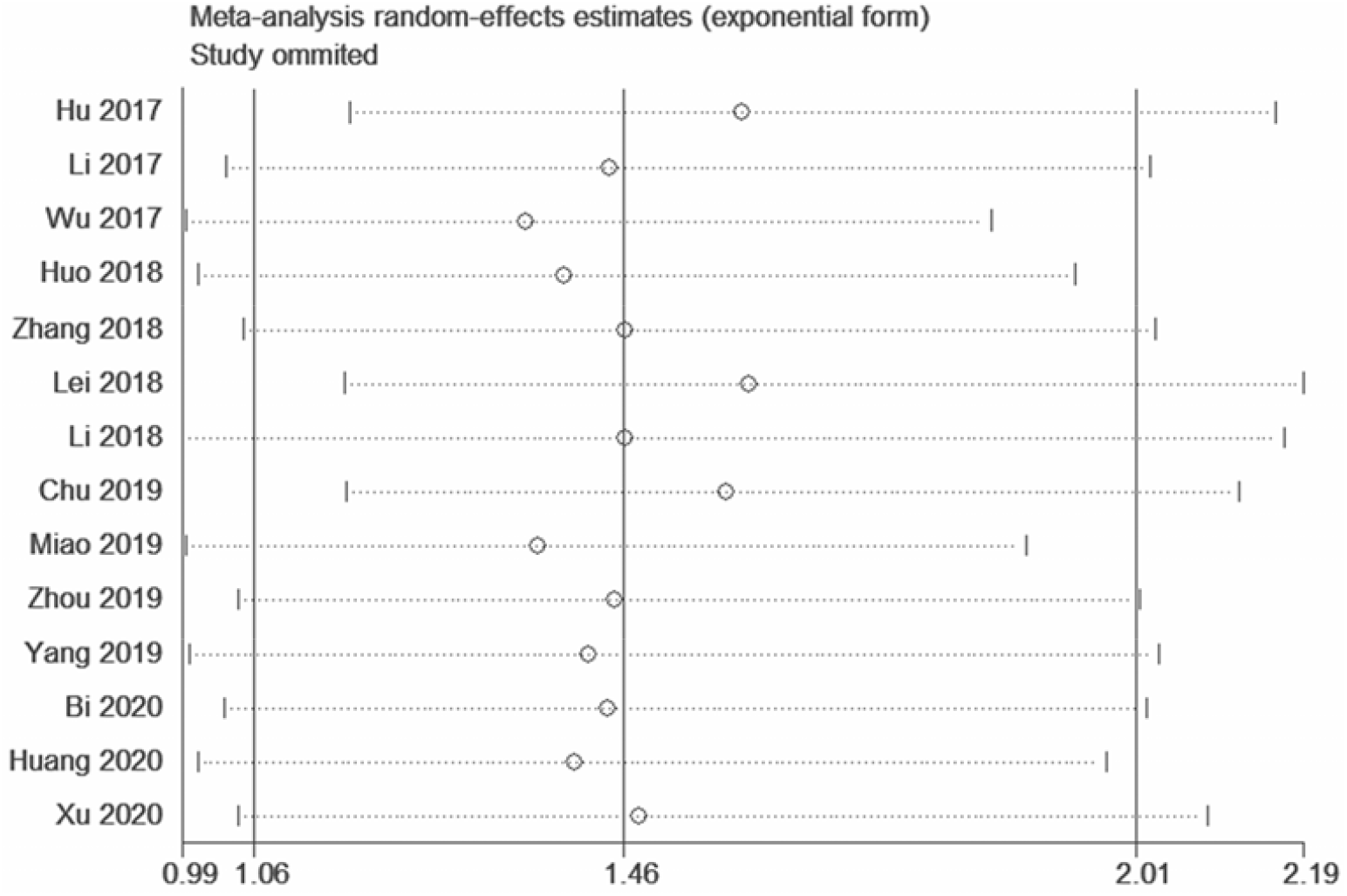
Sensitivity analysis of the relationship between NORAD expression and OS.

There were 2 studies that focused on the correlation of NORAD expression level with DFS. A significant association that high NORAD expression was a risk factor for DFS (HR = 1.74, 95% CI: 1.18-2.57, P = 0.005, Figure 4) was also observed. No heterogeneity was existed among studies (I^2^ = 0.00%). As for RFS, there was only one study reported the association, which indicated that patients with high NORAD expression had poor RFS [33].

**Figure 4.**
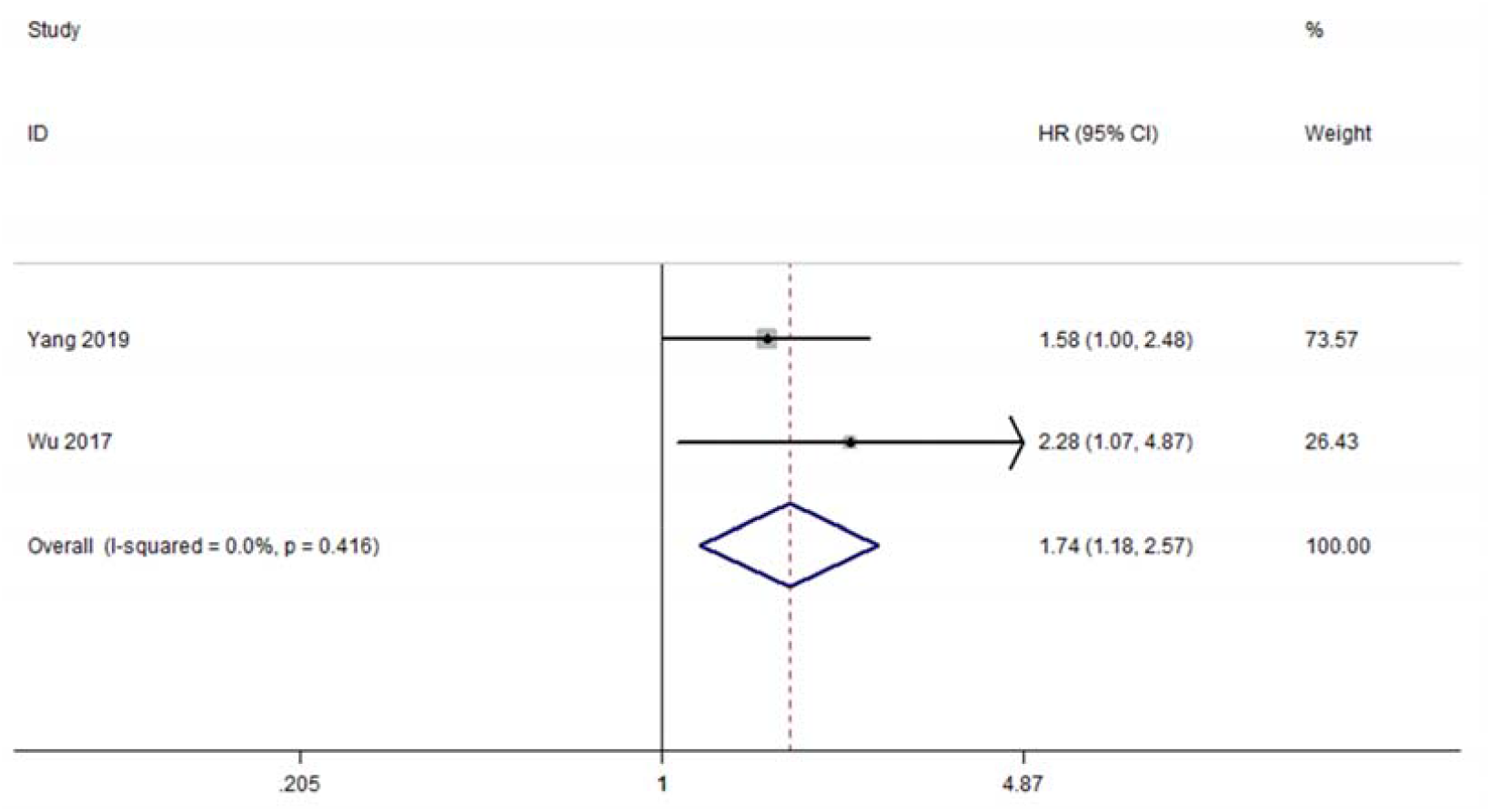
Forest plot for the association between NORAD expression levels and DFS.

### 3.3 Correlation of NORAD expression with clinical characteristics

We further explored the clinicopathological significance of NORAD expression in cancers by pooling available clinical parameters data from included studies. The pooled ORs demonstrated that elevated level of NORAD was significantly correlated with lymph node metastasis (positive vs negative: OR = 2.76, 95% CI: 1.02-7.49, P = 0.046) (Figure 5 a) and T stage (T3+T4 vs T1+T2: OR = 4.91, 95% CI: 1.94-12.44, P = 0.001) (Figure 5 b). However, no significant correlations were observed for age, gender, differentiation, tumor size, distant metastasis, tumor number, liver cirrhosis and vascular invasion (Supplementary Figure S1).

**Figure 5.**
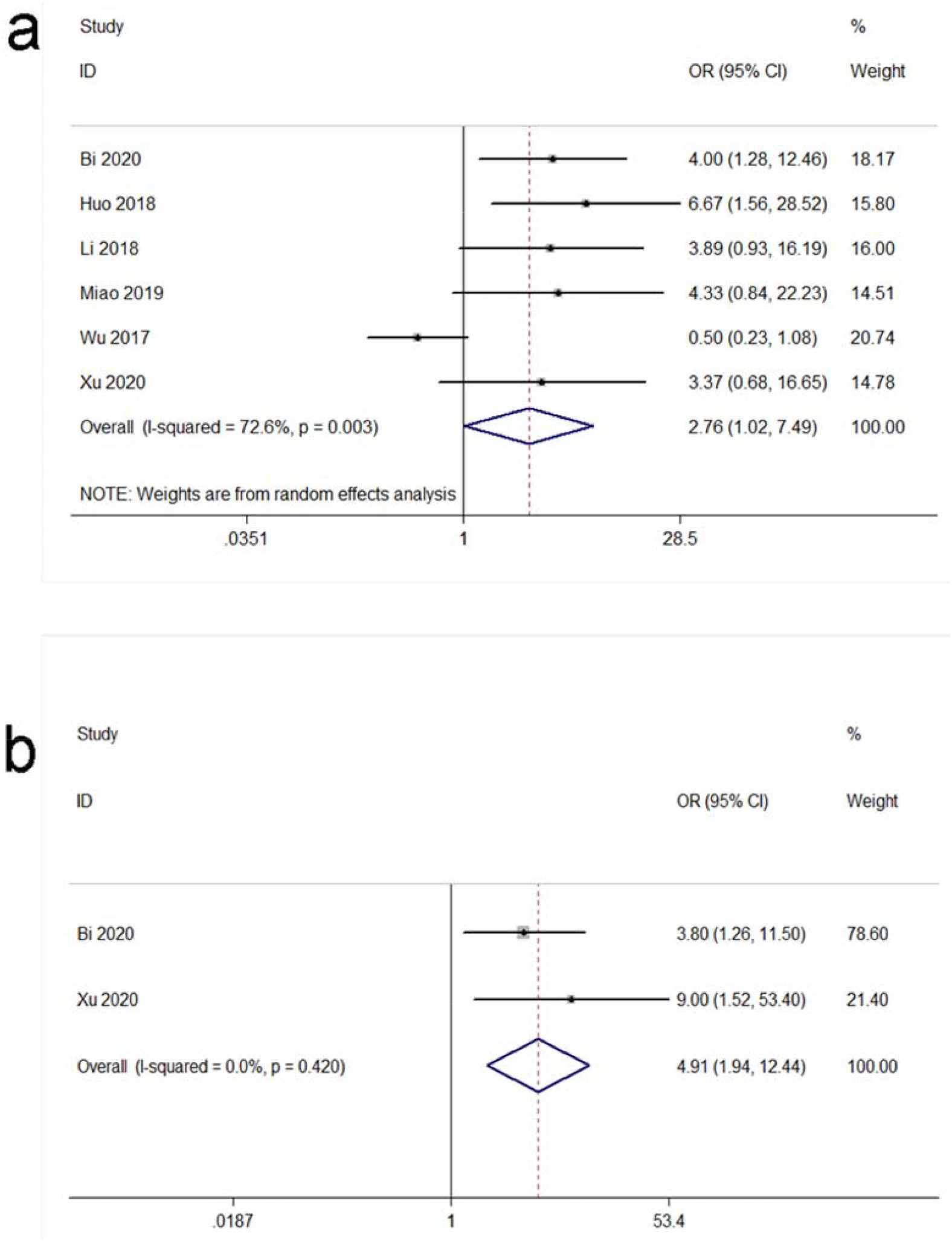
Forest plots for the association between NORAD expression with clinicopathological parameters in cancer patients. (a) lymph node metastasis; (b) T stage.

### 3.4 Publication bias

We utilized Begg’s funnel plots and Egger’s test to detect potential publication bias. Symmetric funnel plots for OS showed that there was no potential publication bias (Figure 6). Moreover, Egger’s test also showed similar results (OS: P = 0.941). Additionally, given the numbers of included articles for DFS, RFS and clinical features were relatively small, analysis for publication bias was not applicable in these groups.

**Figure 6.**
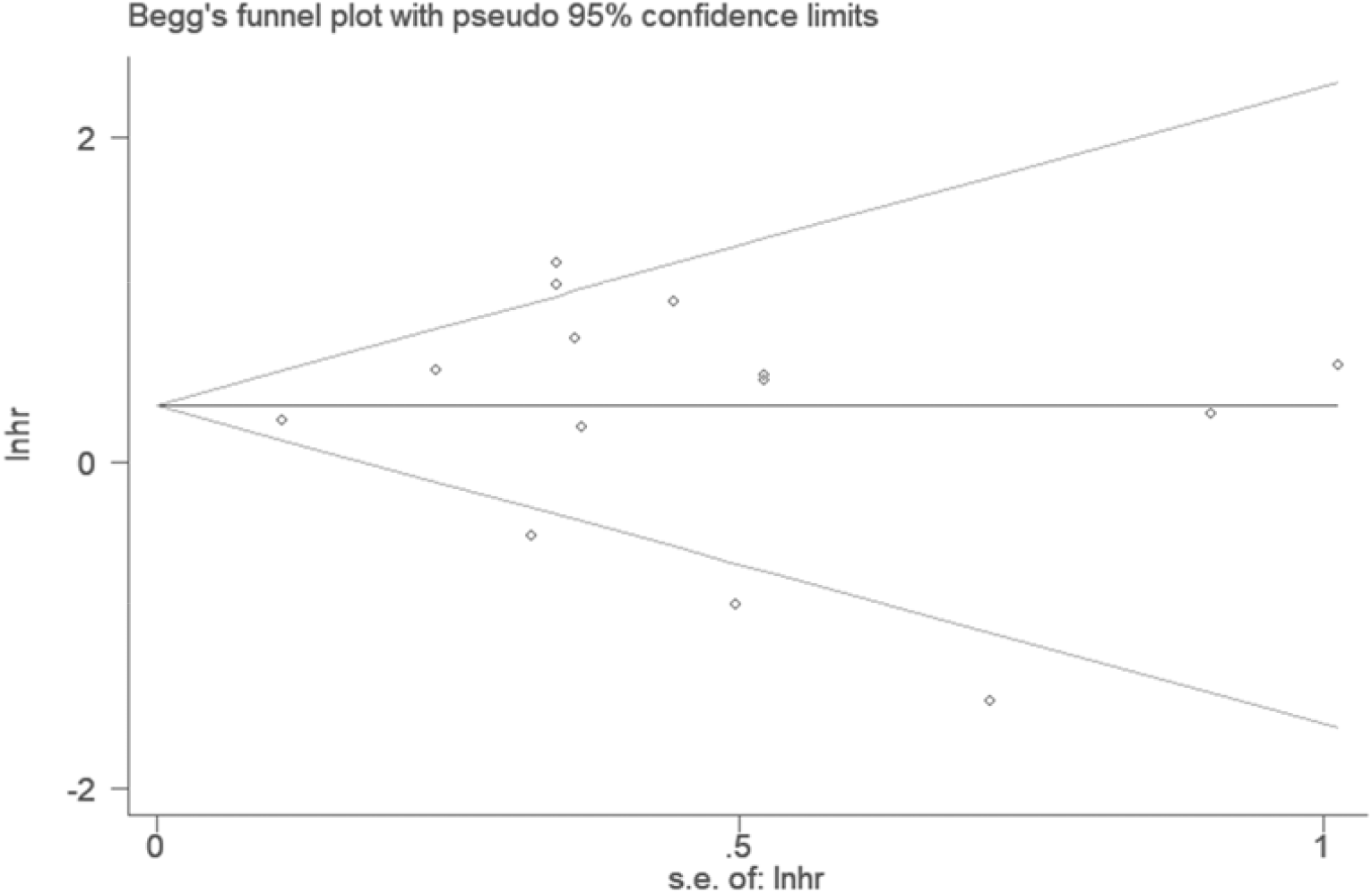
Funnel plots (Begg’s method) of potential publication bias for OS.

### 3.5 Validation of the prognostic role of NORAD in human cancers

Based on GEPIA database, it was showed that NORAD was overexpressed in different cancers compared with corresponding normal tissues (Supplementary Figure S2). Besides, survival analysis indicated that high NORAD expression was significantly related to worse OS and DFS in diverse cancers (Supplementary Figure S3).

## 4. Discussion

This is so far the first comprehensive meta-analysis to assess the prognostic and clinical significance of NORAD in diverse cancers, which indicated that elevated level of NORAD was significantly correlated with poor prognosis, especially for NSCLC, GC, CC, BC and ESCC according to subgroup analysis based on caner types. Moreover, the pooled results demonstrated that overexpression of NORAD was also relevant to lymph node metastasis and T stage. Additionally, the results of present meta-analysis were further strengthened and complemented by the survival information obtained from GEPIA online database. Taken together, these findings provided evidence that lncRNA NORAD served as a promising cancer-related candidate for prognosis and potential targeted therapy.

As one of the promising cancer-related biomarkers for prognosis, the mechanisms of NORAD involved in cancer progression were extensively investigated. Several studies demonstrated that high NORAD expression acted as a competitive endogenous RNA (ceRNA) by sponging miRNA to regulate target genes of miRNA, and thereby exerted its oncogenic role [19, 21,26,37-42]. For instance, it was demonstrated that NORAD positively regulated JunB expression via serving as a ceRNA to sponge miR-615-3p, and then contributed to ESCC progression [38]. Furthermore, NORAD was suggested to function as a ceRNA by upregulating E2F transcription factor 1 (E2F1) via sponging miR-136-5p to promote NSCLC progression [39]. Moreover, NORAD was also found to participate in other cancers progression by regulating miR-590-3p/SIP1, miR-656-3p/AKT1, miR-433/PAK4, miR-26a-5p/CKS2 and miR-608/ FOXO6 axes [19,26,37,41,42]. In addition, other mechanisms of NORAD as an oncogene were to activate different signaling pathways, including RhoA/ROCK1 and TGF-β/RUNX2 signaling pathway, and then facilitate cancer proliferation and invasion [21,40]. In order to verify the correlations between NORAD and these relevant target genes, correlation analysis was conducted by using GEPIA, which demonstrated that NORAD was positively related with PAK4 (R = 0.15, P = 0.044), E2F1 (R = 0.19, P = 4.2e-09), FOXO6 (R = 0.32, P = 2.3e-11) and AKT1 (R = 0.33, P = 0) (Supplementary Figure S4). These abovementioned mechanisms and findings further supported our findings that overexpression of NORAD was a risk factor for OS in cancers.

It was interesting that subgroup analysis based on follow-up time and sample size showed that overexpression of NORAD was significantly correlated with OS in studies with follow-up ≥ 60 months and sample size > 60, but not in studies with follow-up < 60 months or sample size ≤ 60. These data indicated that larger sample size with longer follow-up time were able to provide more convincing evidence, while relatively small sample size or shorter follow-up time would lead to deficient statistical power of the correlation results and mask the actual association between NORAD expression and prognosis in cancers. In this regard, the present meta-analysis is especially necessary as combined results based on the whole current study profile would provide more accurate information of the association between NORAD and cancer outcomes.

In addition, subgroup analysis based on cancer types also suggested that low NORAD expression was significantly related to poor OS in GBM. This finding was reasonable since it was previously reported that elevated level of NORAD suppressed GBM growth and migration via sponging miR-190a-3p to upregulate PTEN expression [25]. However, as only one included study focused on GBM so far with relatively small sample size, this result still needs to be further verified and more well-designed clinical investigations are warranted for further clarification of this issue. Anyway, these subgroup analysis data indicated not only that NORAD might act as a tumor suppressor in GBM but also that the roles of NORAD might be tissue-specific and varied with cancer types. As cancer is a kind of heterogeneous disease, this discrepancy may arise from the heterogeneous molecular profiles embedded in the diverse cancer types [43].

Recent studies also explored the underlying mechanisms for the roles of NORAD in cancer migration and invasion, which reported that NORAD exerted its metastatic properties mainly by inducing epithelial-mesenchymal transition (EMT) in different ways [19,33,40,44,45]. For instance, it was shown that high NORAD expression triggered EMT by upregulating RhoA expression to accelerate pancreatic cancer metastasis [33]. Besides, NORAD induced EMT by modulating the expression of EMT-related genes (i.e., Snail, ZEB1 and E-cadherin), and promoted cancer metastatic properties [19,40]. Additionally, it was also shown that NORAD promoted EMT process by interacting with miR-202-5p or miR-199a-3p, and then contributed to cancer metastasis [44,45]. Consistent with these mechanisms, the current meta-analysis indicated that high expression of NORAD was relevant to advanced T stage and lymph node metastasis, further demonstrating that NORAD was a promising candidate for cancer prognosis and a potential therapeutic target.

However, there were several limitations should be noted. Firstly, the sample size and type of cancers included in the present study were still limited, which may lead to deficient statistical power of our results. Secondly, some of the HRs and 95% CI were extracted indirectly, which might lead to inaccurate data. Thirdly, the application of our findings in other ethnicities and regions may be limited because all patients included in the current meta-analysis were from China. These limitations should be considered in future investigations.

In summary, our results demonstrated that lncRNA NORAD was a risk factor for survival outcomes and clinicopathological parameters in cancer patients. These findings provided a comprehensive insight that lncRNA NORAD may be a promising candidate for prognosis prediction and potential therapeutic target in diverse cancers although further investigations with large scale sample size in different ethnicities still should be conducted for further confirmation of this issue.

## Data Availability

No datasets were generated or analyzed during the current study.

## Declarations

### Compliance with ethical standards

#### Ethics approval

No ethical approval was required for this analysis because all results and analyses were based on previous ethically-approved studies and this article does not contain any studies with human participants or animals performed by any of the authors. This meta-analysis was performed in accordance with the Preferred Reporting Items for Systematic Reviews and Meta-Analyses (PRISMA) statement.

#### Informed consent

No informed consent was required for this analysis because all results and analyses were based on previous ethically-approved studies.

#### Consent to participate

This article does not contain any studies with human participants.

#### Consent to publish

Not applicable.

## Authors’ contributions

Study conceive and design [Qin Yang, Zheng Zhang, Yuan-Yuan Gong, Gong-Hao He], acquisition of subjects and data (Qin Yang, Zheng Zhang, Yuan-Yuan Gong, Gong-Hao He), analysis and interpretation of data (Qin Yang, Zheng Zhang, Yuan-Yuan Gong, Zhi-Ran Li, Hua-Zhu Zhang, Gong-Hao He), and preparation of manuscript (Qin Yang, Zheng Zhang, Yuan-Yuan Gong,, Gong-Hao He).

---

**Table 1**. Characteristics of articles included meta-analysis.

**Table 2**. Stratified analyses for the association of NORAD expression with OS.

**Supplementary Table S1**. The search strategies for PubMed.

**Supplementary Figure S1**. Forest plots for the association between NORAD expression with clinicopathological parameters in cancer patients. (a) age; (b) gender; (c) differentiation; (d) tumor size; (e)distant metastasis; (f) tumor number; (g) liver cirrhosis; (h) vascular invasion.

**Supplementary Figure S2**. The expression levels of NORAD in seven kinds of cancer tissues and normal tissues.

**Supplementary Figure S3**. Survival curves of NORAD in various types of cancers from TCGA database. (a) The overall survival curve of patients with diverse cancers; (b) The disease-free survival curve of patients with diverse cancers.

**Supplementary Figure S4**. Correlation analysis between gene expression levels. (a) Correlation between NORAD and PAK4 expression; (b) Correlation between NORAD and E2F1 expression; (c) Correlation between NORAD and FOXO6 expression; (d) Correlation between NORAD and AKT1 expression.

**PRISMA 2009 Checklist**

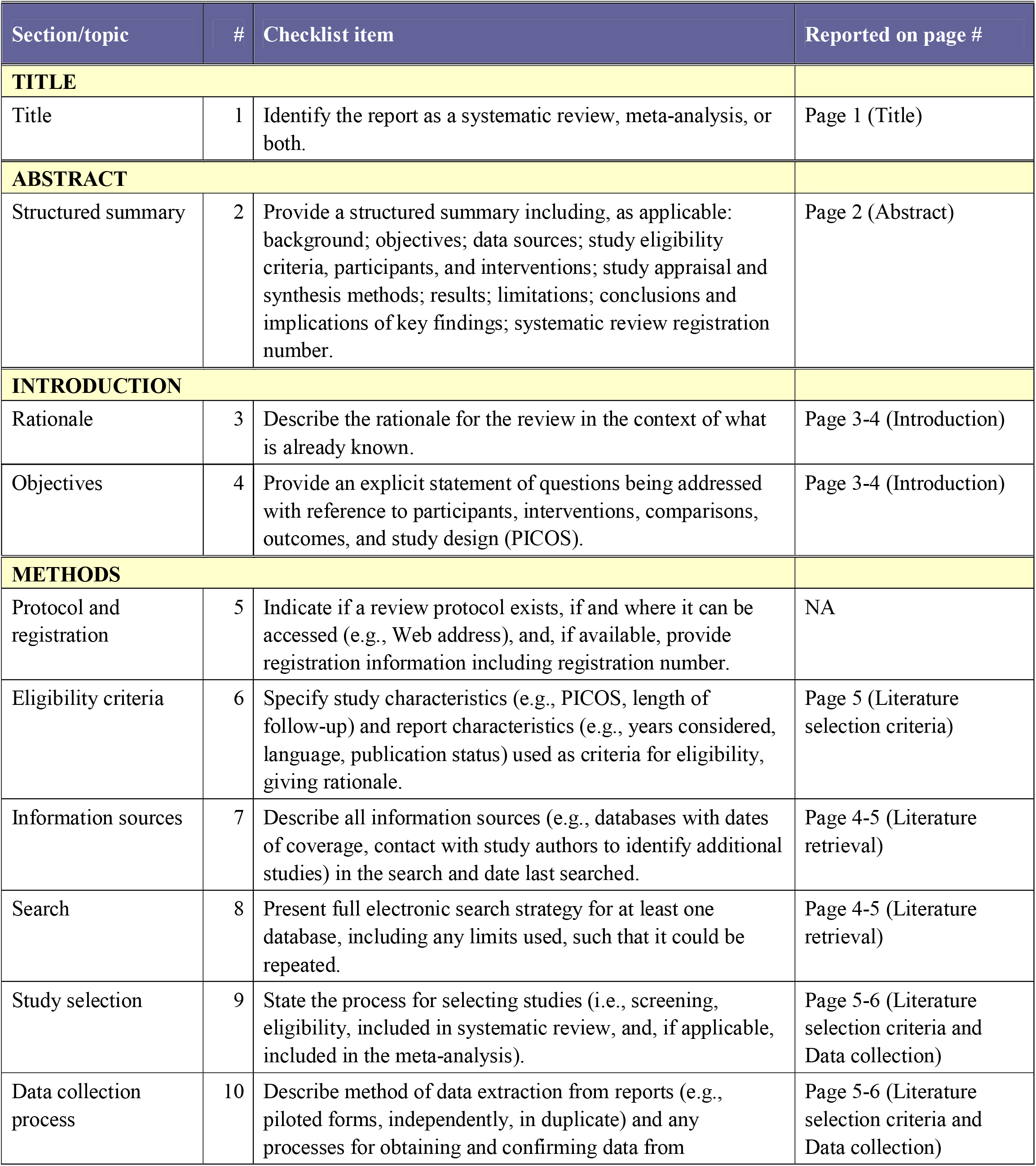
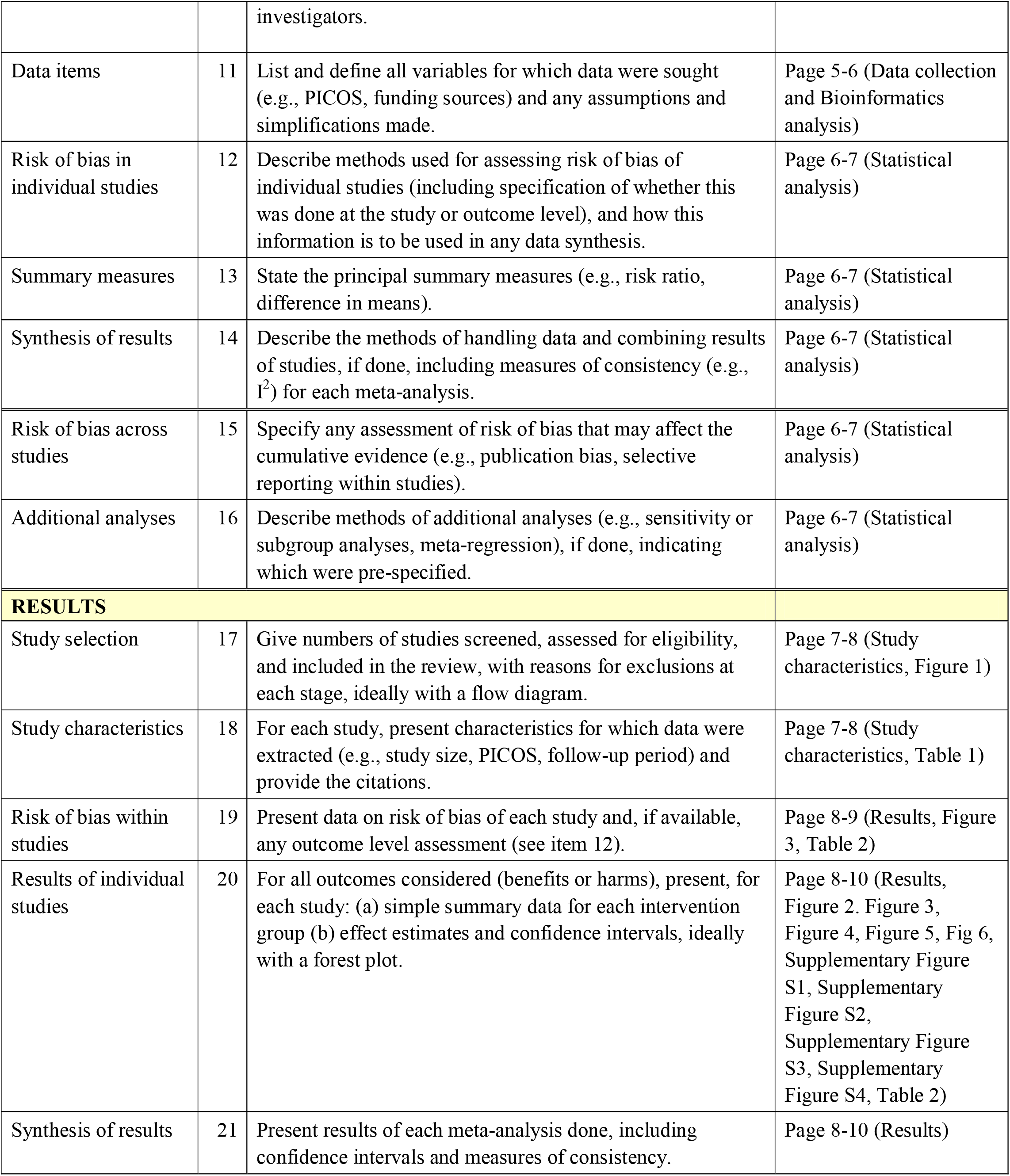
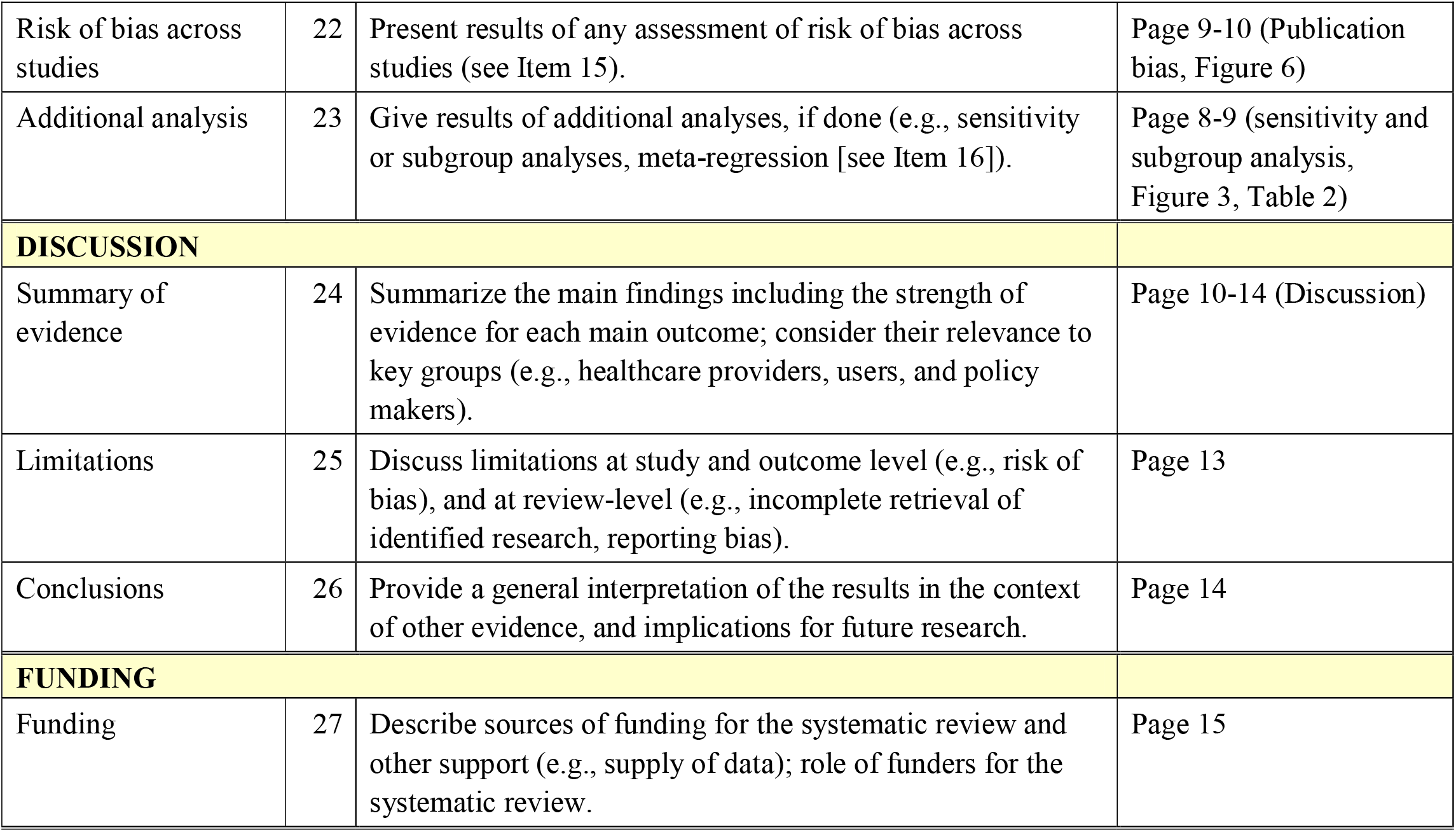

## References

[1] Nair M, Sandhu SS, Sharma AK. Cancer molecular markers: A guide to cancer detection and management. Semin Cancer Biol. 2018; 52:39–55.

[2] Sharma AK. Emerging trends in biomarker discovery: Ease of prognosis and prediction in cancer. Semin Cancer Biol. 2018; 52 (1): iii–iv.

[3] Qi P, Zhou XY, D. X. Circulating long non-coding RNAs in cancer: current status and future perspectives. Mol Cancer. 2016; 15 (1):39.

[4] Miller K D, Nogueira L, Mariotto A B, et al. Cancer Treatment and Survivorship Statistics, 2019. CA Cancer J Clin. 2019; 69(5):363–385.

[5] Sui F, Ji M, Hou P. Long non-coding RNAs in thyroid cancer: Biological functions and clinical significance. Mol Cell Endocrinol. 2018; 469:11–22.

[6] Huang Y, Liu N, Wang J P, et al. Regulatory long non-coding RNA and its functions. J Physiol Biochem. 2012 ;68(4):611–618.

[7] Sánchez Y, Huarte M. Long non-coding RNAs: challenges for diagnosis and therapies. Nucleic Acid Ther. 2013; 23(1):15–20.

[8] Li C H, Chen Y. Targeting long non-coding RNAs in cancers: progress and prospects. Int J Biochem Cell Biol. 2013; 45(8):1895–1910.

[9] Gibb E A, Brown C J, Lam W L. The functional role of long non-coding RNA in human cancers. Mol Cancer. 2011; 10:38.

[10] Zhang H, Chen Z, Wang X, Huang Z, He Z, Chen Y. Long non-coding RNA: a new player in cancer. J Hematol Oncol. 2013; 6:37.

[11] Li J, Xuan Z, Liu C. Long non-coding RNAs and complex human diseases. Int J Mol Sci. 2013; 14(9):18790–18808.

[12] Yao Y, Li J, Wang L. Large intervening non-coding RNA HOTAIR is an indicator of poor prognosis and a therapeutic target in human cancers. Int J Mol Sci. 2014; 15(10):18985–18999.

[13] Cerk S, Schwarzenbacher D, Adiprasito J B, et al. Current Status of Long Non-Coding RNAs in Human Breast Cancer. Int J Mol Sci. 2016;17(9):1485.

[14] Tehrani S S, Karimian A, Parsian H, Majidinia M, Yousefi B. Multiple Functions of Long Non-Coding RNAs in Oxidative Stress, DNA Damage Response and Cancer Progression. J Cell Biochem. 2018; 119(1):223–236.

[15] Silva A, Bullock M, Calin G. The Clinical Relevance of Long Non-Coding RNAs in Cancer. Cancers (Basel). 2015;7(4):2169–2182.

[16] Meseure D, Drak Alsibai K, Nicolas A, Bieche I, Morillon A. Long Noncoding RNAs as New Architects in Cancer Epigenetics, Prognostic Biomarkers, and Potential Therapeutic Targets. Biomed Res Int. 2015; 2015:320214.

[17] Lavorgna G, Vago R, Sarmini M, Montorsi F, Salonia A, Bellone M. Long non-coding RNAs as novel therapeutic targets in cancer. Pharmacol Res. 2016; 110:131–138.

[18] Yang Z, Zhao Y, Lin G, Zhou X, Jiang X, Zhao H. Noncoding RNA activated by DNA damage (NORAD): Biologic function and mechanisms in human cancers. Clin. Chim. Acta. 2019; 489:5–9.

[19] Huo H, Tian J, Wang R, Li Y, Qu C, Wang N. Long non-coding RNA NORAD upregulate SIP1 expression to promote cell proliferation and invasion in cervical cancer. Biomed. Pharmacother. 2018; 106:1454–1460.

[20] Li Q, Li C, Chen J, et al. High expression of long noncoding RNA NORAD indicates a poor prognosis and promotes clinical progression and metastasis in bladder cancer. Urol. Oncol. 2018; 36(6):310.e15-310.e22.

[21] Zhou K, Ou Q, Wang G, Zhang W, Hao Y, Li W. High long non-coding RNA NORAD expression predicts poor prognosis and promotes breast cancer progression by regulating TGF-β pathway. Cancer Cell Int. 2019; 19:63.

[22] Yang X, Cai J B, Peng R, et al. The long noncoding RNA NORAD enhances the TGF-β pathway to promote hepatocellular cancer progression by targeting miR-202-5p. J. Cell. Physiol. 2019; 234(7):12051–12060.

[23] Hu B, Cai H, Zheng R, Yang S, Zhou Z, Tu J. Long non-coding RNA 657 suppresses hepatocellular cancer cell growth by acting as a molecular sponge of miR-106a-5p to regulate PTEN expression. Int. J. Biochem. Cell Biol. 2017; 92:34–42.

[24] Wu X, Lim Z F, Li Z, et al. NORAD Expression Is Associated with Adverse Prognosis in Esophageal Squamous Cell Cancer. Oncol Res Treat. 2017; 40(6):370–374.

[25] Chu L, Yu L, Liu J, et al. Long intergenic non-coding LINC00657 regulates tumorigenesis of glioblastoma by acting as a molecular sponge of miR-190a-3p. Aging (Albany NY). 2019; 11(5):1456–1470.

[26] Miao Z, Guo X, Tian L. The long noncoding RNA NORAD promotes the growth of gastric cancer cells by sponging miR-608. Gene. 2019; 687:116–124.

[27] Xu F Y, Xu X, Hu X D. LINC00657 promotes malignant progression of oral squamous cell cancer via regulating microRNA-150. Eur Rev Med Pharmacol Sci. 2020; 24(5):2482–2490.

[28] Tierney J F, Stewart L A, Ghersi D, Burdett S, Sydes M R. Practical methods for incorporating summary time-to-event data into meta-analysis. Trials. 2007; 8:16.

[29] Stang A. Critical evaluation of the Newcastle-Ottawa scale for the assessment of the quality of nonrandomized studies in meta-analyses. Eur J Epidemiol. 2010; 25(9):603–5.

[30] Tang Z, Li C, Kang B, Gao G, Li C, Zhang Z. GEPIA: a web server for cancer and normal gene expression profiling and interactive analyses. Nucleic Acids Res. 2017; 45(W1):W98–W102.

[31] Toy H I, Okmen D, Kontou P I, Georgakilas A G, Pavlopoulou A. HOTAIR as a Prognostic Predictor for Diverse Human Cancers: A Meta-and Bioinformatics Analysis. Cancers (Basel). 2019; 11(6). Doi: 10.3390/cancers11060778.

[32] Higgins J P, Thompson S G, Deeks J J, Altman D G. Measuring inconsistency in meta-analyses. BMJ. 2003; 327(7414):557–60.

[33] Li H, Wang X, Wen C, et al. Long noncoding RNA NORAD, a novel competing endogenous RNA, enhances the hypoxia-induced epithelial-mesenchymal transition to promote metastasis in pancreatic cancer. Mol. Cancer. 2017; 16(1):169.

[34] Lei Y, Wang Y H, Wang X F, Bai J. LINC00657 promotes the development of colon cancer by activating PI3K/AKT pathway. Eur Rev Med Pharmacol Sci. 2018; 22(19):6315–6323.

[35] Zhang J, Li X Y, Hu P, Ding Y S. LncRNA NORAD contributes to colorectal cancer progression by inhibition of miR-202-5p. Oncol. Res. 2018; Vol. 26, pp. 1411–1418.

[36] Huang Q, Xing S, Peng A, Yu Z. NORAD accelerates chemo-resistance of non-small-cell lung cancer via targeting at miR-129-1-3p/SOX4 axis. Biosci. Rep. 2020; 40(1): BSR20193489.

[37] Bi S, Wang Y, Feng H, Li Q. Long noncoding RNA LINC00657 enhances the malignancy of pancreatic ductal adenocancer by acting as a competing endogenous RNA on microRNA-433 to increase PAK4 expression. Cell Cycle. 2020; 19(7):801–816.

[38] Sun Y, Wang J, Pan S, et al. LINC00657 played oncogenic roles in esophageal squamous cell cancer by targeting miR-615-3p and JunB. Biomed. Pharmacother. 2018; 108:316–324.

[39] Gao W, Weng T, Wang L, et al. Long non[coding RNA NORAD promotes cell proliferation and glycolysis in non[small cell lung cancer by acting as a sponge for miR[136[5p. Mol Med Rep. 2019; 19(6):5397–5405.

[40] Yu S Y, Peng H, Zhu Q, et al, Xiang H G. Silencing the long noncoding RNA NORAD inhibits gastric cancer cell proliferation and invasion by the RhoA/ROCK1 pathway. Eur Rev Med Pharmacol Sci. 2019; 23(9):3760–3770.

[41] Chen T, Qin S, Gu Y, Pan H, Bian D. Long non-coding RNA NORAD promotes the occurrence and development of non-small cell lung cancer by adsorbing MiR-656-3p. Mol Genet Genomic Med. 2019; 7(8):e757.

[42] Zhang X M, Wang J, Liu Z L, Liu H, Cheng Y F, Wang T. LINC00657/miR-26a-5p/CKS2 ceRNA network promotes the growth of esophageal cancer cells via the MDM2/p53/Bcl2/Bax pathway. Biosci Rep. 2020; 40(6):BSR20200525.

[43] Wu Y, Ding M, Wei S, et al. The prognostic value of long noncoding RNA ZEB1-AS1 on clinical outcomes in human cancer. J Cancer. 2018; 9(20):3690–3698.

[44] He H, Yang H, Liu D, Pei R. LncRNA NORAD promotes thyroid cancer progression through targeting miR-202-5p. Am J Transl Res. 2019; 11(1):290–299.

[45] Xu C, Zhu L X, Sun D M, Yao H, Han D X. Regulatory mechanism of lncRNA NORAD on proliferation and invasion of ovarian cancer cells through miR-199a-3p. Eur Rev Med Pharmacol Sci. 2020; 24(4):1672–1681.

